# Serosurvey in two rural areas evidences recent and previously undetected WNV and SLEV circulation in Costa Rica

**DOI:** 10.1101/2021.09.09.21263313

**Authors:** Marta Piche-Ovares, Mario Romero-Vega, Diana Vargas-González, Daniel Barrantes-Murillo, Claudio Soto-Garita, Jennifer Francisco-Llamas, Alejandro Alfaro-Alarcón, Carlos Jiménez, Eugenia Corrales-Aguilar

## Abstract

West Nile virus (WNV) and Saint Louis encephalitis virus (SLEV) share similar virus transmission cycles that involve birds as amplifiers and mosquitoes as vectors. Mammals, including humans, are dead-end-hosts that may be asymptomatic or develop more severe symptoms. Costa Rica is a hyperendemic country for several flaviviruses such as Dengue (DENV) and Zika (ZIKV) and previous research showed limited and restricted SLEV and WNV circulation in horses, sloths, and monkeys. Nevertheless, actual seroprevalence and high transmission areas are not yet identified. To determine putative WNV and SLEV circulation, we sampled peri-domestic and domestic animals, humans, wild birds, and mosquitoes in rural households located in two DENV and ZIKV hyperendemic regions during the rainy and dry seasons of 2017-2018 and conducted PRNT assays for serology and RT-PCR for virus detection. At Cuajiniquil, serological evidence of WNV and SLEV was found in equines, humans, chickens, and wild birds. Also, 5 seroconversion events were recorded for WNV (2 equine), SLEV (1 human), and DENV-1 (2 humans). At Talamanca, a lack of WNV circulation was found, but evidence of SLEV circulation was recorded in equines, humans, and wild birds. No evidence of active viral infection was found in any mosquitoes or wild bird samples. This seroconversion evidence supports the active and recent circulation of SLEV and WNV in these two regions. This study provides clear-cut evidence of WNV and SLEV circulation and should be considered by the health and epidemiology authorities for future prevention and differential diagnostics.

**Author summary:** Mosquitoes serve as vectors for the transmission of infectious diseases such as WNV and SLEV. The natural virus cycle of these viruses is maintained between birds and mosquitoes. Yet, humans and horses are dead end-hosts and can develop severe diseases such as encephalitis. We aimed to elucidate if WNV or SLEV were silently circulating in two regions of the country that historically report numerous cases of other arboviruses such as Dengue and Zika. Eight households were sampled at each region twice during the rainy (high number of arbovirus related infections are reported) and dry season (lower number of infections reported) to record seroconversion events. Serum samples from different species were analyzed using serology and virus presence was detected through molecular methods for wild bird and mosquito pools samples. We found serological evidence of WNV and SLEV infection in horses, humans, wild birds, and chicken samples, but did not detect actual virus in any tissue or mosquito sample. Taken together, our result shows the active but silent circulation of those viruses at both sampling sites. Action to include these arboviruses into diagnostics and public health measures must be taken.

## Introduction

*Flavivirus* genus is composed of single-stranded RNA viruses, which include Dengue virus (DENV), Saint Louis encephalitis virus (SLEV), Zika virus (ZIKV), West Nile virus (WNV), and Yellow Fever virus (YFV) [1, 2]. All of them are arboviruses that cause mosquito-borne diseases throughout the Americas and are responsible for thousands of deaths and hospitalizations every year [3, 4]. Many factors are recognized to contribute to the wide dissemination of those viruses, for instance poorly planned urbanization, geographical expansion of vectors, changing environmental conditions and deforestation [5-7].

The biological cycles of these viruses include a wide variety of susceptible species such as humans, rodents, horses, and non-human primates [8]. The clinical presentation of acute flavivirus infections in humans and animals ranges from mild illness (e.g., asymptomatic infection (50%-80%)) or self-limiting febrile episodes to severe and life-threatening diseases (hemorrhagic fever, shock syndrome, encephalitis, congenital defects) [9-11].

WNV and SLEV belong to the Japanese encephalitis sero-complex [12]. They are neurotropic flaviviruses that cause encephalitis, seizure disorders, and paralysis in humans and equines [10, 13-15]. These viruses are maintained in sylvatic cycles that use birds as amplifiers and mosquitoes of the *Culex* L. complex as vectors [16, 17]. Many species have been associated as primary vectors or moderate vectors in the United States (*Culex tarsalis, Cx. pipiens, Cx restuans, Cx. quinquefasciatus, Cx. stigmatosoma, and Cx. nigripalpus*) [16, 18, 19]. Also, migratory birds can serve as dispersal vectors when they move seasonally and stop at different sites during their journey, establishing possible dispersal events [20]. Mammals, such as equines and humans, serve as dead-end hosts because of the low-level of viremia produced after infection [16, 21]. Severe cases can develop with high fever, neurological dysfunction, altered consciousness, encephalitis, or meningoencephalitis [14, 22].

WNV was introduced to North America from Israel in 1999, causing a widespread outbreak in equines, humans, and wild birds [23]. Following the first report of the virus in the USA, sporadic cases in humans were also reported through the continent (Canada, the Caribbean, and South America) [19, 24]. In Central America, massive avian deaths have not been yet reported, however serological evidence of the circulation of the virus in domestic and wildlife animals has been detected in El Salvador and Guatemala [25-27]. In North America, SLEV has been reported since 1930 [28]. More recently, in 2005 in Argentina, 9 deaths were associated with the virus [29]. In addition, Brazil and Argentina have reported sporadic cases of the disease in people that present mild symptoms like DENV disease, causing a misdiagnosis of the causal agent [30-32].

In Costa Rica, serological evidence of the circulation of WNV was found in equines from Guanacaste with a prevalence between 18-28% in 2004 [33]. In 2009, the first clinical case of WNV was reported in a horse also from Guanacaste, and since then annually new equine cases are reported, especially in the lowlands of the country at the rainy season [34, 35]. However, no bird mortality or human cases associate with the virus are so far recorded. Also, the country lacks information regarding which mosquitoes could be the possible vectors of WNV and SLEV. However, some potential vectors are found present in the country as *Cx. quinquefasciatus, Cx. thriambus, Cx. nigripalpus* [36]. For SLEV, the only serological study was done by Medlin et al. 2016 in sloths (*Choloepus hoffmanni* and *Bradypus variegatus*) from the Caribbean region and antibodies against SLEV and WNV were found in those samples [37].

Costa Rica is endemic for other Flavivirus such as DENV and ZIKV [38, 39]. Molecular epidemiology shows the circulation of DENV 1-3 in humans and molecular and serological evidence of DENV-4 circulation was found in wild animal samples [40, 41]. The co-circulation of different Flavivirus adds complexity in the diagnosis because of significant cross-reactivity and similarities in the undifferentiated fever-like initial symptoms. [42]. The National Health Service of Costa Rica does not consider SLEV and WNV in their routinely diagnostic panel of arbovirus disease, therefore, their epidemiology, and/or local presence in the human population is still poorly studied. The continuous monitoring in endemic areas such as our country but also other tropical areas is crucial to evaluate the risk of transmission to humans and animals [42, 43]. An early detection and timely reporting are fundamental to evaluate the risk of transmission. Surveillance based on a regular sampling of equines, chickens as sentinels and domestic birds has demonstrated a good sensitivity in different countries [42, 43].

The present study aims to evidence the silent circulation of WNV and SLEV in two rural areas of Costa Rica that are hyperendemic for other Flaviviruses such as DENV and ZIKV. Humans, wild birds, equines, and mosquito samples were analyzed to better understand if the viral cycle was present in those areas. We sampled during the rainy and dry seasons of 2017-2018 and conducted PRNT assays for serology and RT-PCR for virus detection. Here we report several seroconversion events in different species, but no evidence of active viral infection was found in any mosquitoes or bird samples. This seroconversion evidence supports the active and recent though silent circulation of SLEV and WNV in these two regions. This information must be taken in account by the health and epidemiology authorities to act for future prevention and differential diagnostics.

## Methods

### Study area

The study was conducted in two regions of Costa Rica where previous flaviviruses infections (DENV, ZIKV and WNV) were officially recorded by Ministerio de Salud de Costa Rica (National Health Service) and the Servicio de Salud Animal (Animal Health Service) [34, 44, 45]. The sampling process was performed during the rainy and dry seasons of 2017 and 2018. The first site was Cuajiniquil, located at the Pacific coast (10°15’06” N, 85° 41’07” O) in the province of Guanacaste in the northwest part of Costa Rica (Fig 1, A). The second site was Talamanca (9°37′14.99″ N, 82 50′39.98″ O) located at the South Caribbean (Fig 2, A).

**Fig 1:**
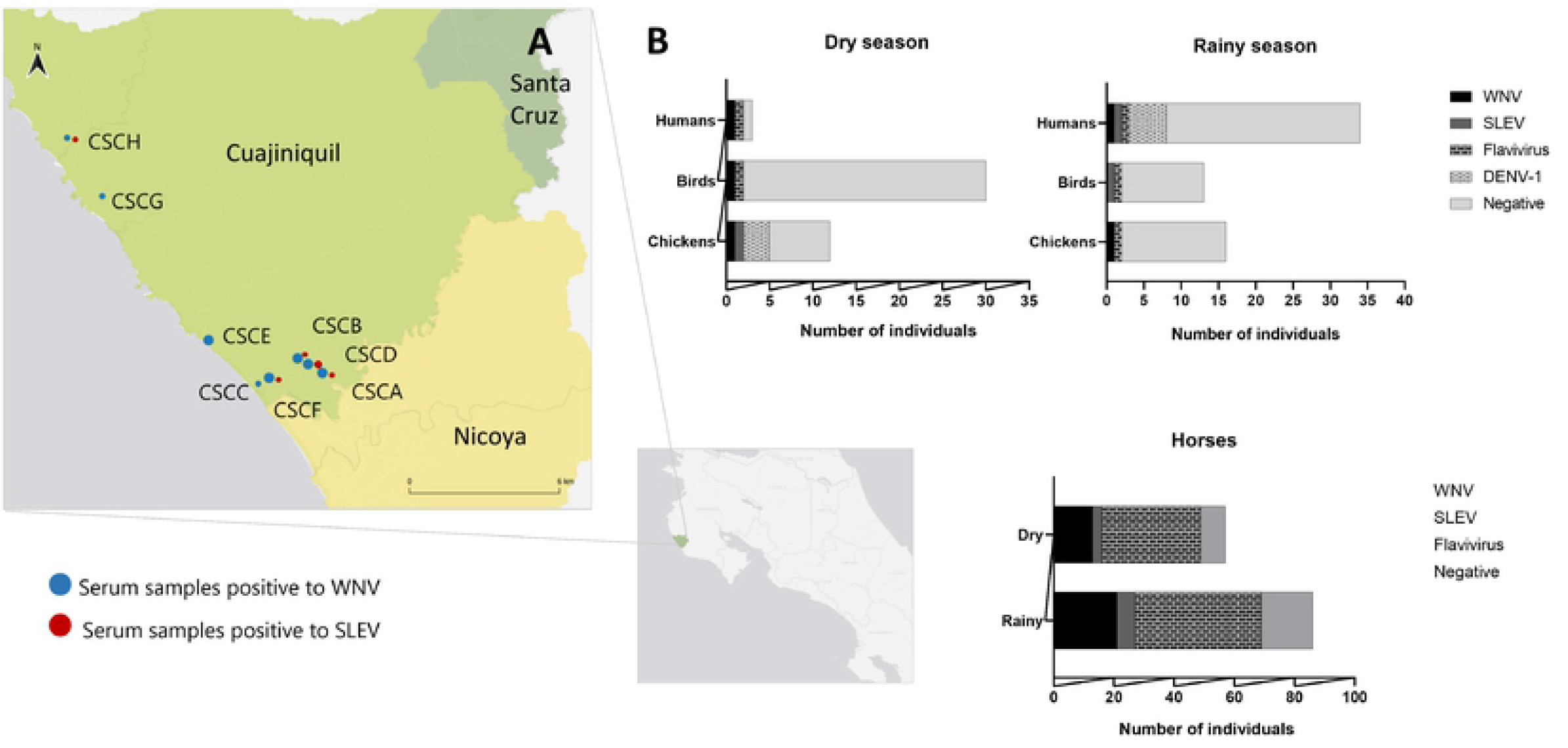
Neutralizing antibodies detected against different Flaviviruses in Cuajiniquil, Costa Rica during the rainy and dry season. A: Geographic distribution of households positive for antibodies of WNV (blue dots) and SLEV (red dots). At each household at least one individual was positive against WNV B: Number of individuals (chickens, wild birds, horses, and humans) positive against different Flaviviruses during the rainy and dry season.

**Fig 2:**
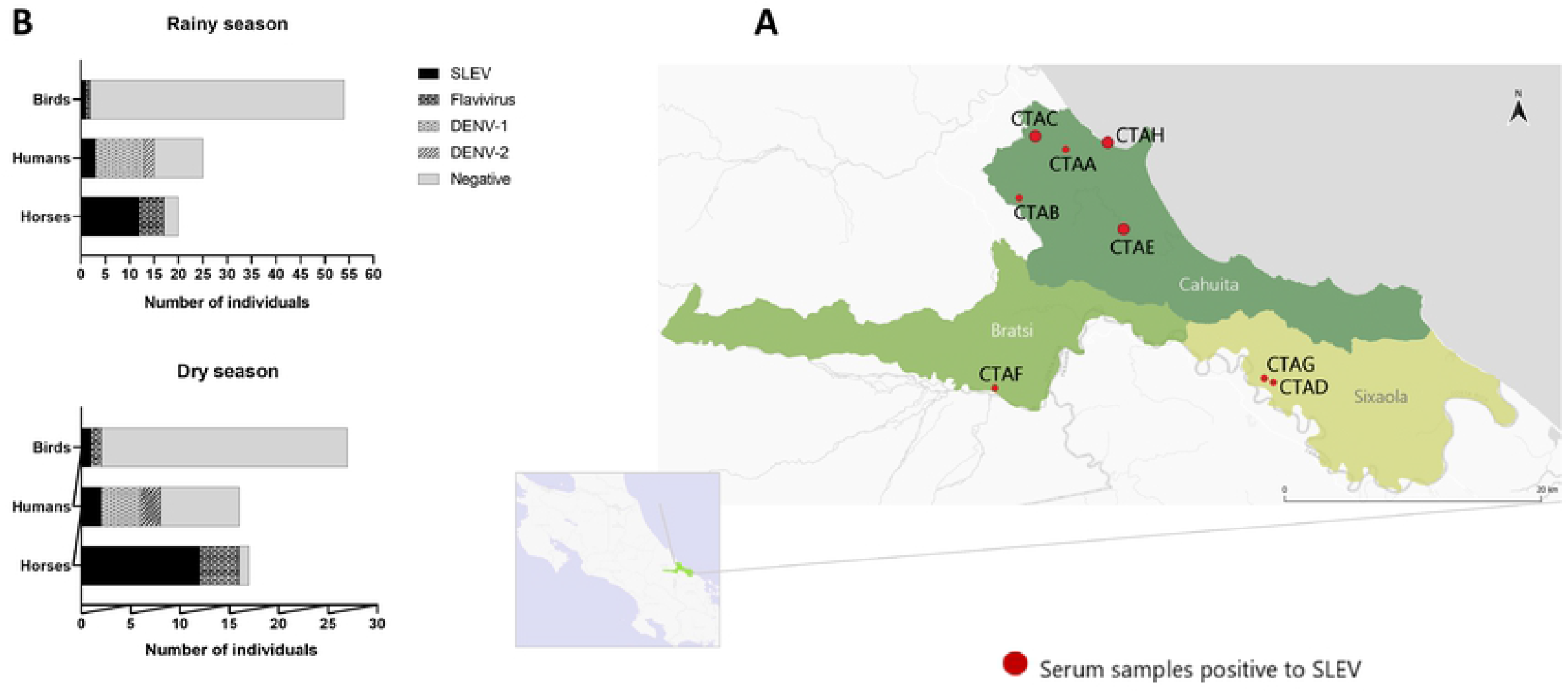
Neutralizing antibodies detected against different Flaviviruses in the Caribbean Coast of Costa Rica during the rainy and dry seasons. A: Geographic distribution of households positive for antibodies of SLEV. At each household at least one individual was positive against SLEV. No evidence of antibodies against WNV was found.

At each site, 8 households were chosen for sampling and serum samples from equines, humans, chickens, and wild birds were taken. At the same time, wild birds were captured using mist-nets and were identify using morphological examination by referring to published identification keys. Then a complete post-mortem analysis was performed. The criteria employed to define the sites for the survey were: (i) presence of at least an unvaccinated for WNV equine, (ii) a forest patch near the household, and (iii) that the household inhabitants were willing to participate and signed the informed consent. Two sampling processes were done at each site: during the rainy and the dry season. The objective was to record putative seroconversion events.

### Sampling and classification of wild birds and mosquitoes, sampling of equines and humans

Birds were captured using mist-nets positioned at two sites (forest and peridomiciliary) in each household. At least, five birds per household were taxonomically identified (S1 text) and then euthanized by an intramuscular anesthesia overdose (ketamine 10 mg/kg + xylazine 1 mg/kg) [46, 47]. Blood sample was taken and stored at 4 °C until arrived at the laboratory where it was stored at - 70 °C for later analysis. Then complete post-mortem analysis and histopathological analysis were performed. Additionally, samples of most organs were aseptically collected (heart, lung, liver, spleen, intestine, kidney, brain, reproductive tract, eye, and proventriculus) and conserved in ARN later® (Thermo Scientific, cat AM0721). Also, a pool of organs was collected in ARN later® for Rt-PCR positivity initial screening.

Field sampling of mosquitoes was done in parallel. Encephalitis vector survey (EVS) traps (BioQuip Products Inc., California, USA) baited with CO_2_ were placed during 12-16 hours in four different locations in each household: inside, peridomiciliary, barn, and forest. Mosquitoes were collected the next morning and transferred to the field lab on ice. A taxonomical identification to species level was done using published keys and compiled in S2 table [36, 48]. Mosquitoes were grouped according to the collection site and species (maximum 20 species per pool). Gravid females were analyzed individually to determine their blood preference.

Blood samples from equines were taken by puncture of the jugular vein, only animals older than 6 months were sampled. Gender, age, breed, and travel history were recorded. Chickens (*Gallus gallus*) samples were taken from the wing vein. The human sample was taken from peripheral venipuncture. Whole blood was centrifuged, and serum was store -20 °C for serological analysis.

### Serological Screening by plaque reduction neutralization tests (PRNT)

Flavivirus exposure was evaluated in sera obtained from horses, humans, domestic chickens, and wild birds by plaque reduction neutralization test (PRNT), considered the gold standard for determining Flavivirus antibodies [49, 50]. For PRNT analysis, different flavivirus-envelope-protein-expressing yellow fever chimeric viruses donated by the CDC were used, except for ZIKV, in which an ATCC reference strain was used [51-53]. Serum samples were heat-inactivated at 56°C for 30 minutes. Then, they were used for an initial screening against WNV and SLEV using a 1:10 dilution [50, 54]. Briefly, samples were diluted 1:5 in MEM with 2% of FBS and mixed with an equal volume of each virus to an estimated end result of 10 UFP/well. The virus-antibodies mix was incubated 1 hour at 37°C in a 5% CO_2_ atmosphere, then a 100 µl volume was inoculated into a VERO (ATCC® CCL-81™) cells monolayer previously seeded in 48 well-plate and incubated for an hour. Then it was removed and 500 µl of MEM with 2% of FBS and 1% of carboxymethylcellulose were added. After 5 days of incubation, plates were fixed with formalin (3.7%) during an hour and stained with crystal violet (1%). Sera that resulted in a 90% of neutralization relative to the average of the viral control (no sera), were considered WNV or SLEV reactive. Due to smaller volumes of sera, wild bird and chicken samples were tested in a 96 well-plate format using a similar protocol and fixated at 3 days [54].

Samples from humans and equines that were considered reactive (90% reduction of foci) were tested in a serial two-fold dilution that ranged from 1:20-1:1280 against WNV (YFV 17D/WNV Flamingo 383-99), DENV 1-4 (YFV 17D/DENV-1 PUO 359, YFV 17D/DENV-2 218, YFV 17D/DENV-3 PaH881/88, YFV 17D/DENV-4 1228), ZIKV (ATCC® VR-748), SLEV (YFV 17D/SLEV CorAn 9124), and YF (YFV 17D) in similar conditions as the previously described protocol. Wild birds and chicken serum samples were only tested against WNV and SLEV because of the limited sera volume. A plaque reduction of ≥90% was considered positive, with the titer measurement as the highest serum dilution showing ≥90% of plaque relative to the average of the viral control. A 4-fold difference in titer between the different flaviviruses was required for unequivocally classifying that serum sample as specifically neutralizing that particular flavivirus. In case that a 4-fold dilution difference was not reached, the serum was classified as flavivirus positive.

### RT-PCR in wild birds and mosquito samples

Viral ARN was extracted from avian tissue (pool of organs) and mosquito pools using the TRIzol ® (Ambion, 15596018) method according to the manufacturer’s instructions. Reverse transcription was done using Revert Aid First Strand cDNA Synthesis Kit (Thermo Scientific cat K1622) with random hexamers primers. A negative (water) and positive control (GAPDH) were included, total ARN of the sample were measure using a NanoDrop™ 2000 (Thermo Scientific, ND-2000).

Firstly, a semi-nested PCR was performed using Flavivirus genus specific primers localized in the NS5 following this protocol previously described [55]. A positive control (YFV 17D) and negative control (water) were included. PCR products were analyzed and quantified using QIAxcel DNA screening gel (Qiagen, 929554), a 220 pb band was expected [55]. Positive samples from the nested PCR were purified using ExoSAP-IT™ (Applied Biosystems, 78201) following manufacturer instructions. Then, a Sanger sequencing of both strands was done by Macrogen Inc. (Seoul, South Korea). The resulting sequence was compared with entries in GenBank database using the nucleotide basic alignment search tool (BLASTn) (https://blast.ncbi.nlm.nih.gov/Blast.cgi) and MEGA X software [56].

### Mosquito blood meal preferences

To determine the bloodmeal preference, gravid females were taxonomically identified and processed individually. Mosquitoes were macerated in a 1.5 mL tube and DNA-RNA was extracted using NucleoSpin® TriPrep (740966.50, Macherey-Nagel). ARN that was obtained from these samples was analyzed against flaviviruses as previously described [55].

Bloodmeal preference was determined using a set of primers for cytochrome oxidase subunit I (COI), following the protocol of Townzen et al. 2008 [57]. PCR products were purified using ExoSAP-IT™ (Applied Biosystems, 78201) and subjected to nucleotide sequencing with forward and reverse primers at Macrogen Inc (Seoul, South Korea). The sequence was compared with entries in GenBank database using the nucleotide basic alignment search tool (BLASTn) (https://blast.ncbi.nlm.nih.gov/Blast.cgi) and MEGA X software [56].

### Statistical analysis

Statistical analysis was conducted using R v3.6.2. A chi-square test was used to assess the correlation between gender and WNV and SLEV seropositivity. A Spearman rank correlation coefficient (*Rs*) was used to correlate WNV seropositivity and age and Flavivirus positivity α= 0.005.

### Ethical statement

The study and associated protocols were designed based on national ethical legislation and approved by the Institutional Committee of Care and Use of Animals of the University of Costa Rica (CICUA-042-17), Committee of Biodiversity of the University of Costa Rica (VI-2994-2017), National System of Conservation Areas (SINAC): Tempisque Conservation Area (Oficio-ACT-PIM-070-17), La Amistad-Caribe Conservation Area (M-PC-SINAC-PNI-ACLAC-047-2018). The survey did not involve endangered or protected species.

After signing of an informed consent previously approved by the University of Costa Rica’s Ethic Scientific Committee (CEC or IRB in English) (CEC-VI-4050-2017), a blood sample was taken from humans for serology analysis.

## Results

### Several flaviviruses co-circulate in each sampled region, WNV and SLEV as silent ones

Eight households were sampled at each arbovirus-hyperendemic region during the rainy and dry seasons of 2017-2018. At each household, serum samples from equines, humans, chickens, and wild birds were taken. At the same time, wild birds were captured using mist-nets and a complete post-mortem analysis was performed.

Serum samples from different species were analyzed using serology by PRNT ≥90% to record seroconversion events. A total of 106 equines, 34 humans, 39 chickens, and 140 wild birds were tested. In Cuajiniquil, Guanacaste (Fig 1, B) during the rainy season, 36 (41.86%) of the equines, 1 (6.25%) human, and 1 (3.45%) chicken were indistinctively positive for neutralizing antibodies against WNV (4-fold dilution of difference) (Table 1 and 2). Also, serological evidence for SLEV was found in 11 (12.79%) equines, 1 (7.69%) wild bird and 1 (3.45%) chicken. This analysis also showed that 5 (31.25%) of the human samples has antibodies against DENV-1 (Table 2). At a later time point and during the dry season, samples were taken from the same individuum (except for wild birds and chickens) to record putative seroconversion events. Five seroconversions were detected: 2 for WNV in a horse, 1 for SLEV and 2 for DENV-1 in humans. Also, 1 (2.86%) wild bird found positive had neutralizing antibodies against WNV (Fig 1, B). No serological evidence of DENV-2,3,4, ZIKV, and YF was found. At this site 15 horses (17.44%), 4 humans (33.33%), 37 chickens (97.43%) and 86 (98.83%) wild birds were negative for all the analyzed viruses. The wild bird species that presented neutralizing antibodies against WNV was identified as *Campylorhynchus rufinucha*, a resident very common species in that area [46].

**Table 1:** Serological characterization of the samples. List of individuals with neutralization antibodies against WNV and SLEV in Cuajiniquil. Two seroconversion events for WNV were recorded.

| Cuajiniquil |  |  |  |  |
| --- | --- | --- | --- | --- |
| Species | Animal identification | Age (years) | PRNT titer |  |
|  |  |  | WNV | SLEV |
| Equines | ESCA2 | 7 | 1:320 | 1:1280 |
|  | ESCA3 | 8 | 1:640 | 1:80 |
|  | ESCA4 | 6 | 1:640 | 1:80 |
|  | ESCA5<sup>a</sup> | 6 | >1:1280 | <1:40 |
|  | ESCA6 | 3 | 1:1280 | 1:40 |
|  | ESCA7 | 6 | 1:20 | 1:160 |
|  | ESCA8 | 2 | >1:1280 | 1:40 |
|  | ESCA9 | 5 | 1:160 | **1:640** |
|  | ESCA12 | 5 | **1:640** | 1:40 |
|  | ESCB2 | 3 | **1:160** | 1:20 |
|  | ESCB3 | 5 | 1:40 | **1:640** |
|  | ESCB4 | 3 | **1:160** | 1:40 |
|  | ESCB7 | 2 | **1:160** | 1:20 |
|  | ESCB8 | 5 | **1:640** | 1:40 |
|  | ESCC2 | 4 | **1:640** | 1:20 |
|  | ESCD1 | 4 | **1:1280** | 1:80 |
|  | ESCD2 | 3 | **1:160** | 1:20 |
|  | ESCD4 | 10 | 1:40 | **>1:1280** |
|  | ESCD5 | 10 | **>1:1280** | <1:40 |
|  | ESCD8 | 8 | **1:640** | 1:20 |
|  | ESCD9 | 10 | **1:160** | <1:40 |
|  | ESCD10 | 2 | **>1:1280** | <1:40 |
|  | ESCD11 | 12 | **1:1280** | <1:40 |
|  | ESCD15 | 22 | **1:320** | 1:80 |
|  | ESCD17 | 3 | **1:640** | <1:40 |
|  | ESCD19 | 2 | **1:640** | <1:40 |
|  | ESCD24 | 1,5 | **1:160** | <1:40 |
|  | ESCD26 | - | **1:1280** | <1:40 |
|  | ESCE1 | 3 | **1:320** | 1:20 |
|  | ESCF1 | 8 | **>1:1280** | 1:320 |
|  | ESCF3 | 12 | **1:640** | 1:160 |
|  | ESCF6 | 10 | 1:80 | **1:320** |
|  | ESCF9 | 10 | **1:160** | 1:40 |
|  | ESCF10 | 8 | **1:160** | <1:40 |
|  | ESCF13 | 5 | **1:640** | 1:40 |
|  | ESCF14 | 12 | **1:640** | 1:160 |
|  | ESCF15 | 4 | **1:320** | 1:20 |
|  | ESCF16 | 2 | 1:40 | **1:640** |
|  | ESCF18 | 10 | **1:320** | 1:20 |
|  | ESCF20 | 6 | **1:160** | <1:40 |
|  | ESCF21 | 8 | **1:320** | 1:40 |
|  | ESCF24 | 5 | 1:40 | **1:160** |
|  | ESCF25 | 8 | 1:20 | **1:320** |
|  | ESCF27 | 5 | 1:640 | **1:5120** |
|  | ESCF28 | 1 | **1:320** | 1:20 |
|  | ESCG2 | 10 | 1:40 | **1:640** |
|  | ESCG6 | 17 | **1:320** | 1:40 |
|  | ESCG7<sup>a</sup> | 16 | **1:640** | 1:80 |
| **Wild birds** | ASCD1 *Turdus grayi* | Ad | 1:10 | **1:80** |
|  | ASCM9 *Campylorhynchus rufinucha* | Ad | **1:80** | >1:20 |
| **Chickens** | GSCA1 | Ad | **1:80** | 1:10 |
|  | GSCB1 | Ad | 1:40 | **>1:640** |

<sup>a</sup> **Seroconversion events, Bold: Positive, Ad: Adult.**

**Table 2:** Neutralization titers in humans against different flaviviruses in the two regions. In Cuajiniquil antibodies against WNV, SLEV and DENV-1 and two seroconversions events, one for SLEV and one for DENV-1, were recorded.

| **Human identification** | **DENV-1** | **DENV-2** | **DENV-3** | **DENV-4** | **WNV** | **YFV** | **SLEV** | **ZIKV** |
| --- | --- | --- | --- | --- | --- | --- | --- | --- |
| **Cuajiniquil** |  |  |  |  |  |  |  |  |
| HSCA1<sup>a</sup> | **1:320** | 1:160 | 1:40 | <1:20 | <1:20 | <1:20 | <1:20 | <1:20 |
| HSCA2 | 1:80 | 1:80 | <1:20 | <1:20 | **>1:640** | <1:20 | 1:20 | <1:20 |
| HSCA3 | **>1:640** | 1:320 | 1:40 | <1:20 | <1:20 | <1:20 | <1:20 | 1:40 |
| HSCC1 | **1:640** | <1:20 | <1:20 | <1:20 | <1:20 | <1:20 | <1:20 | <1:20 |
| HSCD2 | **1:640** | <1:20 | <1:20 | <1:20 | <1:20 | <1:20 | <1:20 | <1:20 |
| HSCD3 | **1:160** | <1:20 | <1:20 | <1:20 | <1:20 | <1:20 | <1:20 | <1:20 |
| HSCF2 | **1:160** | <1:20 | <1:20 | <1:20 | <1:20 | <1:20 | <1:20 | <1:20 |
| HSCH1<sup>a</sup> | <1:20 | <1:20 | <1:20 | <1:20 | <1:20 | <1:20 | **1:320** | <1:20 |
| **Talamanca** |  |  |  |  |  |  |  |  |
| HTAA2 | <1:20 | <1:20 | <1:20 | <1:20 | <1:20 | <1:20 | **<1:160** | <1:20 |
| HTAD1 | **1:640** | **1:640** | <1:20 | <1:20 | <1:20 | <1:20 | 1:20 | <1:20 |
| HTAD2 | **1:160** | <1:20 | <1:20 | <1:20 | <1:20 | <1:20 | <1:20 | <1:20 |
| HTAE1 | 1:40 | 1:80 | 1:40 | 1:20 | <1:20 | <1:20 | **1:640** | <1:20 |
| HTAF1 | **1:160** | <1:20 | <1:20 | <1:20 | <1:20 | <1:20 | <1:20 | <1:20 |
| HTAF2 | **1:640** | <1:20 | <1:20 | <1:20 | <1:20 | <1:20 | <1:20 | <1:20 |
| HTAG1 | <1:20 | <1:20 | <1:20 | <1:20 | <1:20 | <1:20 | **1:160** | <1:20 |
| HTAH1 | **>1:640** | 1:40 | <1:20 | <1:20 | <1:20 | <1:20 | <1:20 | <1:20 |
| HTAH2 | **1:640** | <1:20 | <1:20 | <1:20 | <1:20 | <1:20 | <1:20 | <1:20 |
| HTAH3 | **1:320** | **1:160** | 1:20 | <1:20 | <1:20 | <1:40 | <1:20 | <1:20 |

<sup>a</sup> **Seroconversion events, bold: Positive**

In contrast, we did not detect any serological evidence for WNV in Talamanca (Fig 2, B), but evidence of previous contact with SLEV. Neutralizing antibodies against SLEV were found in 12 (60%) equines, 3 (17.65%) humans, and 2 (2.47%) wild birds (Table 2 and 3). Also, five (29.41%) human samples were positive against DENV-1 and two (11.76%) have neutralizing antibodies against DENV-1 and DENV-2 (Table 2). At this site 8 equines (40%%), 7 humans (41.17%) and 86 wild birds (97.72%) were negative for all flavivirus and no serological evidence for DENV-3, DENV-4, ZIKV, and YF was found. Wild birds with SLEV-neutralizing antibodies were *Empidonax virescens*, a migratory species that migrate from Canada and *Myiozetetes similis*, a resident species of Costa Rica [46].

**Table 3:**
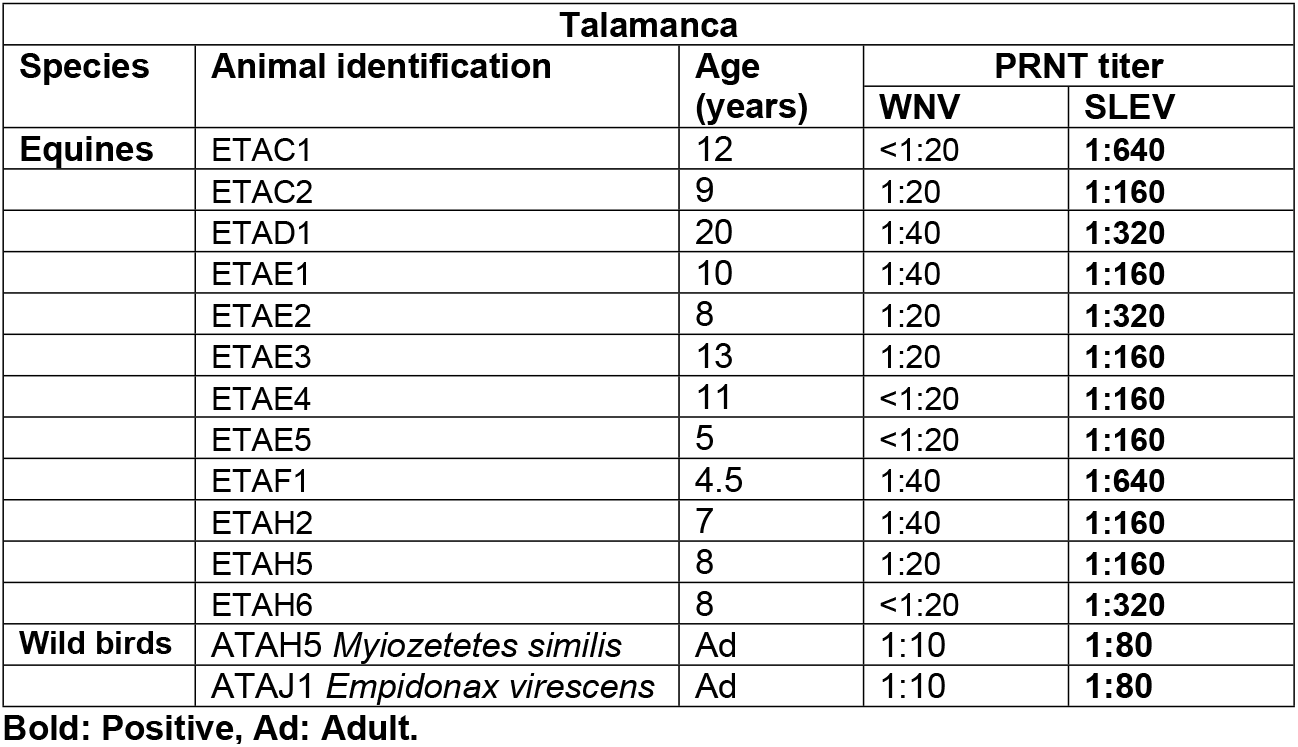
Serological characterization of the samples. List of individuals with neutralization antibodies against SLEV in Talamanca. No evidence of WNV were recorded.

A chi-square test of the equine serum samples was used to assess correlation between gender and WNV or SLEV seropositivity. The results showed no significant correlation between gender and WNV or SLEV seropositivity (X^2^ = 2.2512, df=2, p=0.81).

Some serum samples were classified as Flavivirus positive (n=22, 25.58%) since they reacted with more than one of the viruses tested. These samples were analyzed by a Spearman correlation (*R*_s_) to assess if age and Flavivirus positivity were related. This resulted in a moderate positive correlation (*R*_s_= 0.3625 (*p*=0.05 y *p*<0.01)) (Fig 3). Two main reasons could explain these results, first that animals throughout life have sequential infections by different flavivirus or that other flaviviruses that were not considered in this study are circulating in those areas.

**Fig 3:**
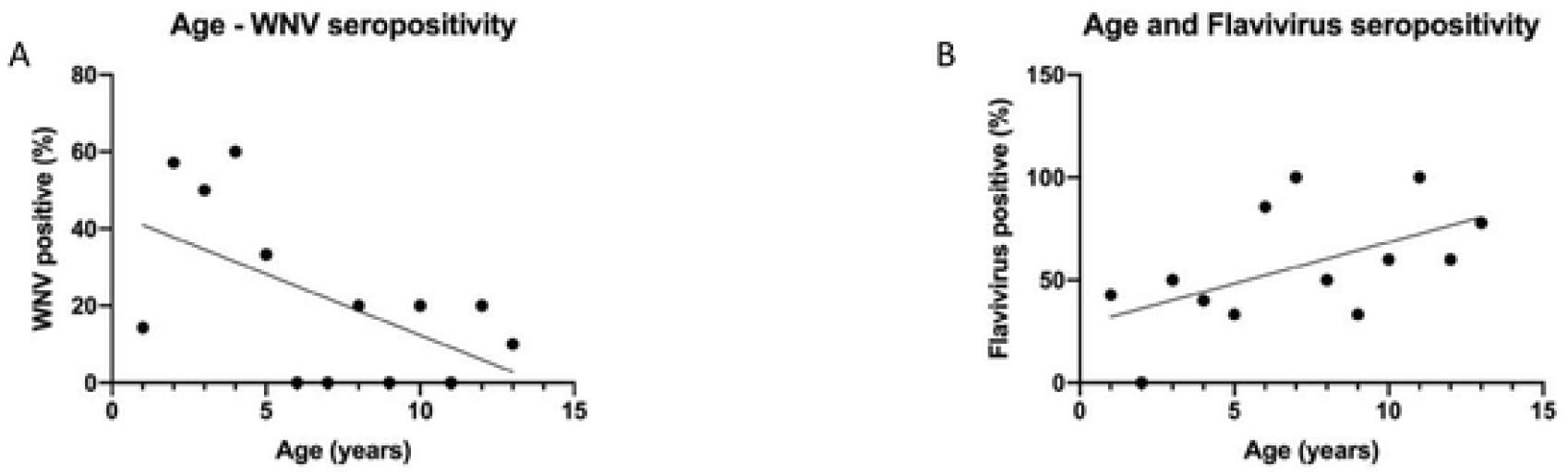
Spearman positive correlation (*R*_s_= 0.3625 (*p*=0.05 y *p*<0.01)) between age and Flavivirus positivity. A: Association between the age and the WNV seropositivity in equines of the Pacific Coast. B: Association between the age and Flavivirus seropositivity in equines of the Pacific Coast.

### No molecular evidence of active virus circulation was found in mosquitoes and wild bird samples

To study the epizootic cycle of these arboviruses, mosquitoes and wild birds were sampled. A total of 140 wild birds were collected during the period of the study. The complete post-mortem and histopathological analysis show no associated lesions to arbovirus infections. In Cuajiniquil area 52 wild birds from 15 different species were captured; 2 species were migratory (S1 table). In Talamanca, 88 wild birds were captured from 29 different species, 6 species were migratory. This area is a very important point of migration from North America to South America [58]. Also, 1373 mosquitoes were captured in 128-night tramps. The most frequent species in the Cuajiniquil area (n=554) were *Deinocerites pseudes* (24.91%, n=138), *Cx. quinquefasciatus* (17.69%) and *Anopheles albimanus* (8.66%). In the area of Talamanca (n=819), the most frequent species were *Cx. quinquefasciatus* (45.91%, n=376), *Cx. coronator* (11.60%, n=95), and *Mansonia titillans* (10.01%, n=82). The complete classification of the mosquitoes according to the location and the sampled season is available in S2 table.

Mosquitoes were grouped according to the collection site and species (maximum 20 species per pool). Gravid females were analyzed individually to determine their blood preference. Mosquito pools (n=164 for Cuajiniquil and n=198 for Talamanca), gravid females (n=32) and wild birds (n=140) were analyzed by a semi-nested RT-PCR using Flavivirus genus specific primers [55]. PCR products were analyzed and quantified using QIAxcel DNA screening gel (Qiagen, 929554). Two mosquito pools from Talamanca were detected positive for flavivirus (Table 3). Positive pools were submitted for nucleotide sequencing and showed 100% of homology to a mosquito flavivirus. In the case of wild bird samples, no positive PCR results were obtained.

**Table 3:** Mosquitoes positive in Flavivirus RT-PCR and its homology.

| Pool identification | Household | Species | Homology | GenBank |
| --- | --- | --- | --- | --- |
| TCC28 | CTAC | *Culex (Melanoconion) sp* | Mosquito flavivirus | MN856966.1 |
| TDC19 | CTAD | *Aedes aegypti* | Aedes flavivirus | MK241496.1 |

### The analyses of mosquito blood meals show a species diversity of feeding sources

Mosquito blood meals preference was analyzed to establish the diversity of blood meals sources and participation of these identified species in the putative virus cycle. Mosquitoes that were classified as gravid females were taxonomically identified using morphological characters and analyzed for their blood meal preference by detection of COI [57]. Twenty-three of the 32 mosquitoes led to positive DNA amplification: from Cuajiiquil *Cx. restrictor* (n=1), *Cx. quinquefasciatus* (n=1), *Anopheles albimanus* (n=1), and *Deinocerites pseudes* (n=1) and from Talamanca: *Cx. quinquefasciatus* (n=9), *Cx. coronator* (n=3), *Cx. (Melanoconion) sp* (n=2), *Psorophora ferox* (n=1), *Cx. pseudostigmatosoma* (n=2), and *Mansonia titillans* (n=1).

After sequencing and blasting analyses, we detected dog (*Canis lupus familiaris*) (n=6, 31.57%), human (*Homo sapiens*) (n=5, 26.31%), equine (*Equus caballus*), (n=4, 1.05%), sheep (*Ovis aries*) (n=4, 21.05%), and wild bird (*Columbina paserina*) (n=1, 5.2%) blood used as feeding source. One sample showed a mixed blood pattern (dog/human).

## Discussion

In this study we detected a silent circulation of WNV and SLEV in two regions of Costa Rica (Cuajiniquil and Talamanca). Nevertheless, we did not detect virus ARN in wild bird organs or mosquito pools. Therefore, an active circulation of those viruses could only be recorded by the seroconversion events that took place at both sampling sites and through neutralizing antibodies found in wild bird samples. Interestingly, our results show simultaneous circulation of several flaviviruses in the sampled areas: WNV, SLEV, and DENV-1 in Cuajiniquil and DENV-1 and DENV-2 in Talamanca. In these areas positive equines against WNV by IgM and human cases of DENV and ZIKV by the National Health authorities have previously been reported [34, 44, 59]. It is tempting to speculate that human infections by WNV and SLEV have possibly been mistaken as DENV and ZIKV symptoms and thus misdiagnosed.

Serological analysis showed that neutralizing antibodies against WNV and SLEV are uniformly distributed in Cuajiniquil. There, in each household at least one of the sampled species had neutralizing antibodies (wild birds, chickens, equines, and/or humans). On the other hand, on the Caribbean side no evidence of previous contact with WNV was recorded but, serological evidence against SLEV was documented in wild birds, horses, and humans, and likewise each household had at least one species positive. Previous studies made in hamsters show that previous immunity with SLEV confers protection against clinical encephalitis and death after been infected with WNV [60]. This could explain the no case report of WNV disease in Talamanca. The strikingly high seropositivity to WNV in Cuajiniquil and SLEV at both regions reveals that these viruses are widely distributed.

We detected 4 wild birds with neutralizing antibodies (3 for SLEV and 1 for WNV) belonging to four different species. Three of them were resident wild birds (*Campylorhynchus rufinucha, Myiozetetes similis, Turdus gray*) suggesting a local contact with the virus. It is tempting to speculate that the virus cycle mosquito-bird-mosquito is thus locally well established. The species *Empidonax virescen* is a migratory species that was captured in Talamanca. This area is one of the most important sites for wild bird migration in the world [58]. During the yearly migration interval from October to November thousands of wild birds fly over here to South America [58, 61]. This migratory behavior could lead to the introduction of wild bird-hosted flaviviruses but may bring even more new strains of SLEV and WNV to our country [20, 62].

WNV and SLEV share common mosquito vectors (*Culex*) and present comparable transmission cycles and clinical signs [15, 18, 63]. At both sampling areas, *Cx. quinquefasciatus* was one of the most abundant mosquito species collected, other species of *Culex* such as *Cx. nigripalpus* also were identified. This species has been proposed as a vector for WNV and SLEV in America [21, 64-66]. The blood meals detected, and the species distribution further confirms the presence of *Culex* species that serve as bridge vectors capable of transmitting WNV between wild birds and end-hosts e.g., humans and equines.

Furthermore, an important percentage of the equines at both sampling sites were classified as “Flavivirus positive”. This means that they present similar neutralizing antibodies titers for more than one flavivirus. This can be explained by two main reasons: i) that animals came in contact early in their lives with one virus and then have contact with other Flavivirus throughout life, thus sequential infections by different flaviviruses, this is supported by the moderate correlation obtained where older animals have more cross-reactivity compared with younger ones; ii) that other flaviviruses not evaluated in this study also co-circulate. For instance, in Brazil, Ecuador and Bolivia antibodies against Rocio Virus and Ileus were reported in equines [67, 68]. This emphasizes the importance of continued monitoring and detection of different flavivirus species in the country.

Costa Rica like the rest of Latin America, lacks information about the seroepidemiology of WNV and SLEV. Our study demonstrates an ongoing circulation of WNV in the region of Cuajiniquil and SLEV. Also, shows the co-circulation of other Flaviviruses such as DENV and ZIKV, and suggests that others flaviviruses could be also silently circulating [39]. Regions with multiple Flaviviruses encounter a significant challenge in the clinical and serological diagnosis. Laboratory testing is crucial for accurate diagnosis because symptoms can overlap. Almost all ELISA kits are not completely devoid from cross-reactions to properly interpret results in these serological assays, and thus procure potential misinterpretation [42]. Molecular diagnosis by qRT-PCR of serum, plasma, and cerebrospinal fluid is of limited value for routine diagnosis, due to low level and short-lived viremia generated by these viruses [42]. The PRNT ≥90% technique is the gold standard for identifying antibodies against different Flaviviruses, but this technique is expensive, needs laboratory facilities and requires careful interpretation. Because of the above, flaviviruses serological diagnosis is a real challenge [42].

Active surveillance for WNV and SLEV must be prioritized and performed in flavivirus-endemic areas in mosquitoes, wild birds, and sentinel chickens to detect the virus before the outburst of disease or outbreaks in equines and humans. Also, WNV and SLEV must be considered as a differential diagnosis in patients suspected for DENV and ZIKV infection. Further studies must be done to establish the national seroprevalence and genotypes that are circulating in the country. Costa Rica as a tropical country has the potential of introduction and establishment of new flaviviruses that could cause a more complex epidemiologic scenario.

As prior studies show that a previous infection with ZIKV or DENV modulates a second infection with a different virus, increasing the probability of symptomatic and severe disease [69]. Therefore, there are still concerns in the cross-reaction between them, their potential immune interplay, and the challenge for a correct diagnostic.

## Data Availability

All data is available

## Acknowledgments

We thank to all the people who allowed us to sample at their homes. We thank the Arbovirus Reference Collection (ARC) from the Centers for Disease Control and Prevention, National Center for Emerging and Zoonotic Infectious Diseases, Division of Vector-Borne Diseases, Arboviral Diseases Branch, Diagnostic and Reference Team, Reference and Reagent Laboratory and Sanofi Pasteur for the chimeric viruses contribution.

## Supporting information

**S1 Table**. List of captured wild birds species in the 2 sites of study in Costa Rica (Cuajiniquil and Talamanca) during 2017-2018

**S2 Table**. List of captured mosquitoes species by household, tramp, and number of individuals per pool.

